# Health-Framed Messaging and Its Impact on Attitudes Toward Daylight Saving Time

**DOI:** 10.1101/2025.10.11.25337370

**Authors:** Meltem Weger, Courtney von Hippel, Frédéric Gachon, Benjamin D. Weger

## Abstract

**Objectives:** This study aimed to examine public attitudes toward daylight saving time (DST) and its perceived health consequences in Australia, where state-level variation in DST adoption provides a unique setting to test whether the exposure to DST-related health risk information influences these attitudes and how individual differences moderate this effect.

**Methods:** In a preregistered, randomized online experiment, Australian adults (n = 499) were assigned to receive either neutral information about DST or DST-related health-risk messaging highlighting its negative health consequences. Participants’ attitudes toward DST, policy preferences, perceived health consequences, pre-existing health awareness, and credibility perceptions were assessed, and sociodemographic characteristics were subsequently collected. Chronotype was assessed using the Micro-Munich ChronoType Questionnaire.

**Results:** Exposure to DST-related health-risk messaging reduced DST policy support and increased its perceived health consequences, effectively eliminating the modest majority in favour of DST in the absence of such messaging. These effects were stronger among individuals with higher pre-existing health awareness. Mediation analyses showed that shifts in overall attitudes toward DST policy were explained by heightened recognition of DST-related health consequences, with this indirect pathway being amplified when the information was perceived as more credible.

**Conclusions:** Collectively, our findings show that favourable views of DST are linked to lower recognition of its health consequences, while credible health-risk messaging reduces support by raising awareness of these risks. This underscores the importance of evidence-based health-risk messaging for informing DST policymaking in Australia and internationally, where DST remains a matter of debate.

## Introduction

Daylight saving time (DST) is a biannual adjustment in which clocks are set forward one hour in spring and back in autumn with the intent to extend evening daylight during warmer months. DST was first introduced in Germany during World War I as an energy-saving measure and is currently observed in about 70 countries [1]. In Australia, DST applies in most jurisdictions, except for Queensland, Western Australia, and the Northern Territory, all of which have observed DST in the past before ultimately opting out [2].

Largely debunked in its ability to reduce energy consumption [3–6], the regular public and political debate about DST in Australia, as elsewhere, now centres on its potential social and economic benefits, such as extended evening daylight for leisure and commercial activities, reduced crime rates and improved traffic safety [7–9]. However, major sleep and chronobiology societies around the world, including the Society for Research on Biological Rhythms [10,11], the European Biological Rhythms Society [11], the British Sleep Society [12], the American Sleep Research Society [13], and the American Academy of Sleep Medicine [14], have recommended abolishing the biannual clock changes associated with DST in favour of permanent standard time. This recommendation is based on a growing body of evidence linking DST to a range of health risks, such as sleep deprivation, impaired cognitive performance, mood disturbances, obesity, cardiovascular events, mental health crises, and work-related injuries [10,15–19].

From a circadian health perspective, DST contributes to circadian misalignment (“social jetlag”), a mismatch of the body’s internal circadian clock with the environment, by abruptly advancing social time (dictated by clocks) without a corresponding shift in the solar light-dark cycle (determined by the sun). Persistent circadian misalignment has been linked with various health problems [17], a reduced life expectancy [20,21], and substantial economic costs [17,22]. Circadian misalignment and its associated health effects particularly affect late chronotypes [23–25], who are individuals with a natural preference to fall asleep and wake later than average. They account for 20-25% of the population, with higher prevalence during adolescence [26]. Shift workers, who constitute roughly 16% of Australia’s workforce [27,28], are also at risk [29–31].

However, such concerns are rarely addressed in public or political debates. Legislation and referenda in the European Union [32] and the United States [33] have pushed to end biannual clock changes. In contrast, Australia, has not followed a consistent path, and assessments of public attitudes toward DST remain limited, relying mainly on historical referendum data (*e.g.,* Queensland 1992 [34], Western Australia 2009 [35]). To date, it remains unclear whether public support for DST reflects informed acceptance or limited awareness of potential health risks, and to what extent public attitude toward DST can be influenced by DST-related health-risk information. Clarifying these gaps is critical, as it can help to design more effective, evidence-based health communication and policy, not only in Australia but also internationally. This is particularly important considering that public views on DST are often shaped by tradition, personal convenience, and anecdotal reasoning [1,17,36,37].

We examined how and to what extent, DST-related health-risk messaging influences public attitudes toward DST, policy preferences, and beliefs about the likelihood and severity of its negative health effects (*i.e.,* perceived health risks). We further hypothesized that the impact of health-risk messaging would not be uniform across individuals but would depend on individual differences such as pre-existing health-awareness, perceived message credibility, and chronotype.

## Participants and Methods

### Ethics

This study was approved by the University of Queensland Ethics Committee under approval number 2025/HE000340. All procedures were carried out in accordance with relevant guidelines and regulations. Informed consent was obtained from all participants prior to their participation in the survey.

### Study participants and data collection

We recruited 500 voluntary participants via the Prolific platform to obtain a broad and diverse sample across Australia. Data were collected online through a Qualtrics survey on 22 April 2025, when standard time was in effect across all Australian states. In this way, we ensured that responses were not influenced by acute effects of DST transitions. Each participant received £1.05 as compensation. To be eligible, participants had to be Australian residents, at least 18 years old, and proficient in English. In total, 499 participants completed the survey, with one excluded for not providing consent. See Table 1 for an overview of the cohort characteristics.

**Table 1.** Demographic, socioeconomic, and geographic characteristics of participants in the control and health-risk conditions. *Note. Baseline characteristics of participants are presented; groups did not differ significantly on these variables*.

| Characteristic | Control<br>(n = 249)<sup>1</sup> | Health-risk<br>(n = 250)<sup>1</sup> | p-value<sup>2</sup> | Stat |
| --- | --- | --- | --- | --- |
| **Age (years)** | 38 (13) | 39 (14) | .448 | t = -0.76 (df = 495.3) |
| **Gender** | | | .524 | $\chi^2 = 1.26$ (df = 2) |
| Male | 119 (48%) | 107 (43%) |  |  |
| Female | 128 (51%) | 141 (56%) |  |  |
| Other | 2 (0.8%) | 2 (0.8%) |  |  |
| **Occupation** | | | .881 | $\chi^2 = 1.18$ (df = 4) |
| Managerial, professional, technical and scientific | 135 (56%) | 138 (57%) |  |  |
| Clerical and secretarial | 40 (16%) | 35 (14%) |  |  |
| Sales and personal service | 30 (12%) | 36 (15%) |  |  |
| Craft and skilled manual | 18 (7.4%) | 15 (6.1%) |  |  |
| Semi-skilled and unskilled manual | 20 (8.2%) | 20 (8.2%) |  |  |
| **Pre-existing health awareness** | 5.20 (0.96) | 5.19 (0.91) | .848 | t = 0.19 (df = 495.6) |
| **Chronotype (MSF<sub>sc</sub>, h)** | 3.53 (1.32) | 3.24 (1.41) | .032 | t = 2.15 (df = 407.9) |
| **Social jetlag (h)** | 0.51 (0.68) | 0.42 (0.68) | .183 | t = 1.33 (df = 430.9) |
| **Sleep duration (workdays, h)** | 7.80 (1.17) | 7.76 (1.15) | .698 | t = 0.39 (df = 491.0) |
| **Sleep duration (free days, h)** | 8.40 (1.19) | 8.29 (1.21) | .325 | t = 0.99 (df = 489.4) |
| **Shift work** | 35 (14%) | 27 (11%) | .334 | $\chi^2 = 0.93$ (df = 1) |
| **State** | | | .447 | $\chi^2 = 5.77$ (df = 6) |
| NSW/ACT | 82 (33%) | 81 (33%) |  |  |
| QLD | 40 (16%) | 50 (20%) |  |  |
| VIC | 70 (28%) | 60 (24%) |  |  |
| SA | 26 (11%) | 19 (7.6%) |  |  |
| WA | 20 (8.1%) | 28 (11%) |  |  |
| TAS | 8 (3.2%) | 11 (4.4%) |  |  |
| NT | 1 (0.4%) | 0 (0%) |  |  |
| **DST status** | | | .109 | $\chi^2 = 2.70$ (df = 1) |
| DST | 186 (75%) | 171 (69%) |  |  |
| No DST | 61 (25%) | 78 (31%) |  |  |
| **Residence** | | | .846 | $\chi^2 = 0.37$ (df = 2) |
| Urban | 183 (74%) | 181 (73%) |  |  |
| Suburban | 54 (22%) | 60 (24%) |  |  |
| Rural | 9 (3.7%) | 8 (3.2%) |  |  |
| **Latitude (°)** | -33.9 (4.3) | -33.7 (4.3) | .544 | t = -0.61 (df = 494.0) |
| **Longitude (°)** | 145 (10) | 145 (11) | .604 | t = 0.52 (df = 487.5) |

<sup>1</sup> Mean (SD); n (%)
<sup>2</sup> Welch Two Sample t-test or Pearson's Chi-squared test.

### Survey structure

The study was preregistered prior to data collection, with hypotheses, primary outcomes, and analytic procedures specified in advance on OSF (https://doi.org/10.17605/OSF.IO/CTXNW), to ensure that the design was hypothesis-driven and that primary outcomes were clearly defined. Participants were first presented with an information sheet explaining that the survey sought to assess attitudes and opinions regarding DST, that there were no right or wrong answers, that responses would remain anonymous, and that withdrawal was possible at any time. Upon providing consent, all participants were provided with general information about the practice of DST and its adoption in Australia before being randomly assigned to one of two groups: a control condition, in which they received general information about the history of DST, or a health-risk condition, which received information on circadian misalignment and documented health consequences of DST, summarised from publicly available sources cited within the message. After the manipulation, all participants completed a survey designed to measure attitudes toward DST, credibility perceptions, policy preferences, and related constructs. All measures were rated on a 7-point Likert scale ranging from 1 (strongly disagree) to 7 (strongly agree). Composite indices were calculated as the mean of available responses, and cases missing responses to all items were coded as missing. Following the assessment of the key constructs, demographics were collected (*e.g.,* sex, age, occupation, geographic location). No identifying information was collected to preserve participant anonymity.

The primary outcome variable was support for DST, assessed in three complementary ways. General attitudes toward DST were measured with 12 items assessing both positive and negative evaluations (*e.g.,* “Overall, I feel positive about daylight saving time”; “Changing the clocks twice a year is inconvenient”). Six items were reverse scored so that higher scores reflected more favourable attitudes toward DST (Cronbach’s α = .94).

Perceived health consequences of DST were measured using three items (*e.g.,* “I believe daylight saving time has negative effects on health”). Items were averaged into a composite, with higher scores indicating stronger endorsement of health risks (Cronbach’s α = .83). Policy preference for DST was measured with two state-specific items. Participants in states observing DST were asked about keeping the policy (*e.g.,* “I support keeping daylight saving time in my state”), while participants in non-DST states were asked about introducing it (*e.g.,* “I support introducing daylight saving time in my state”). The two items were averaged to form a composite index, with higher scores reflecting stronger support for adopting or maintaining DST (Cronbach’s α = .94).

Secondary measures included general health awareness, assessed with six items (*e.g.,* “I pay close attention to health-related news and information”), that were averaged to form a composite score with higher values reflecting greater attentiveness to health-related topics (Cronbach’s α = .87). Perceived credibility of information was measured with four items (*e.g.,* “The information I read about daylight saving time was credible”), averaged to yield a credibility index with higher scores indicating greater perceived credibility (Cronbach’s α = .88). Both messages were perceived as reasonably credible (control message M = 5.20, SD = 0.88; health-risk message M = 4.94, SD = 0.98), although the control message was rated as slightly more credible [t(491.4) = 3.11, *p* = .002)].

Sleep-wake habits and chronotype were assessed using the validated Micro-Munich ChronoType Questionnaire (µMCTQ; [38]), which captured chronotype as well as related parameters including social jetlag and sleep duration (*e.g.,* “I normally fall asleep/wake up at:”, “I have been a shift- or night-worker in the past three months”).

### Data analysis

All analyses were conducted in R (v4.4.0). We applied data-quality checks to ensure valid responses. Manipulation check items assessed message content recall and credibility of the key element of their assigned condition. The majority of participants responded correctly [236 of 249 (control condition) and 232 of 250 (health-risk condition)]. For confirmatory analyses, we compared primary outcome variables across conditions in both the full sample and in a subset including only participants who passed the manipulation check. Results were substantively unchanged across both approaches. Therefore, we retained the full sample for the reported analyses.

Data from the µMCTQ section of the survey were processed using the R package mctq [39] to derive standard sleep-wake metrics: average sleep duration on workdays and free days, chronotype, and relative social jetlag. Chronotype was operationalised as the mid-sleep point on free days corrected for sleep debt (MSF_sc_). This validated measure indicates whether an individual is more morning-typed (lower MSF_sc_ values, corresponding to earlier sleep timing) or more evening-typed (higher MSF_sc_ values, corresponding to later sleep timing). Relative social jetlag was defined as the difference between midsleep on free days and midsleep on workdays. Positive values reflect a later midsleep on free days when compared to workdays (*i.e.,* delayed sleep on free days), whereas negative values indicate an earlier midsleep on free days (*i.e.,* advanced sleep on free days). Larger absolute values indicate a greater misalignment between biological rhythms and social/work schedules. Input times were normalised to a 24-h format, correcting common entry errors. This included ambiguous time entries such as “12:00”, 4-digit times without separators and missing am/pm. Implausible sleep durations (<3 h or >14 h) were flagged, and ambiguous sleep onsets were shifted by +12 h if doing so restored a plausible duration (6 h - 12 h). Nonsensical entries were set to NA. Latitude, longitude, state, and residence (urban, suburban, rural) were retrieved with participants’ postcodes using data derived from the Geocoded National Address File [40]. States were also classified according to DST status (DST-observing *vs.* non-observing).

For analyses involving gender, participants who selected “non-binary” or “prefer not to say” [n = 2 (control message), n=2 (health-risk message)] were excluded due to the very small subgroup size, which precluded meaningful statistical comparison. Analyses proceeded in several stages. We first ensured that participants in the control and health-risk conditions did not differ systematically in sociodemographic or chronobiological characteristics using chi-square tests for categorical measures and Welch’s t-tests for continuous measures (Table 1). To identify determinants of DST attitudes independent of health-risk messaging, we used multiple regression analyses only in the control condition. Both univariate models (examining each predictor separately) and a multiple variate model (including all predictors simultaneously) were tested, incorporating demographic, geographic, and chronobiological predictors (Table 2). As an exploratory step, we visualised associations between key outcome variables and assessed their correlations using Spearman’s rho. Specifically, we examined the relationships between DST policy preferences and attitudes towards DST, perceived health consequences and policy preferences, and perceived health consequences and attitudes towards DST (Fig. 1 G-I). Next, one-way ANOVAs tested for differences between the control and health-risk message conditions on the main dependent variables (general attitudes towards DST, DST policy preferences, and perceived health consequences). To assess individual differences in message effects, we applied Hayes’ PROCESS-style moderation models [41] to test whether pre-existing health awareness, chronotype, social jetlag, or current DST policy in the state of residence moderated the relationship between condition and attitudes. Significant interactions were probed with simple slopes (Table 3). Finally, moderated mediation analyses (*i.e.,* model 7; [41]) tested whether perceived health consequences mediated the effect of condition on DST attitudes, and whether this indirect effect was moderated by perceived message credibility. Indirect effects were estimated using bias-corrected bootstrapped confidence intervals (5,000 resamples). We ran models unadjusted and adjusted for covariates (age, chronotype, relative social jetlag, DST status, residence), with no substantive differences observed between specifications. All tests were two-tailed with α = 0.05, and false discovery rate (FDR) correction was applied where indicated.

**Fig. 1.**
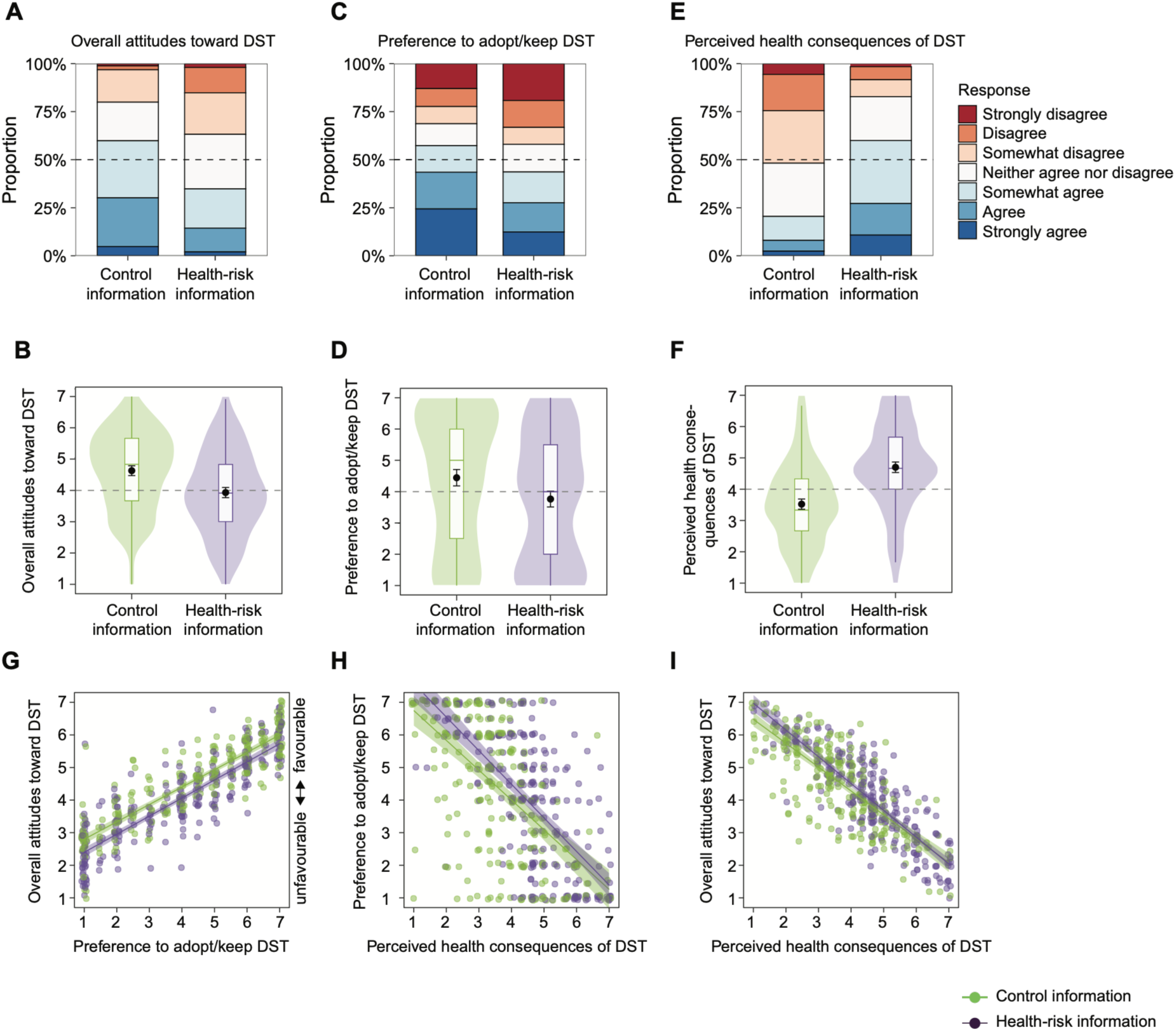
Modest positive attitudes toward DST and support for DST policy are associated with lower perceptions of health consequences. (A-F) Participants were assigned to a control information or health-risk information condition. Panel A, C, and E show percentage distributions across a 7-point Likert scale (1 = strongly disagree, 7 = strongly agree) for overall attitudes toward DST (A), support for adopting/keeping a DST policy (C), and perceived health consequences of DST (E). Panel B, D, and F show the corresponding score distributions as violin plots with embedded Tukey boxplots (median line; box = IQR; whiskers = ±1.5×IQR). Black points mark the mean with 95% CIs. The dashed horizontal line denotes the scale midpoint. (G-I) Spearman rank correlations showing a positive correlation between overall attitudes toward DST and support for adopting/keeping DST policy (G), and inverse correlations between perceived DST-related health consequences and overall attitudes toward DST (H) and support for adopting/keeping DST policy (I). Lines indicate least-squares fits with 95% CIs; points are individual respondents within the control (green) and health-risk message condition (purple) groups.

**Table 2.** Individual difference factors associated with attitudes toward DST in the control group.

| Predictor | Full model |  |  |  | Univariate |  |  |  |  |
| --- | --- | --- | --- | --- | --- | --- | --- | --- | --- |
| | *F*(df<sub>1</sub>, df<sub>2</sub>) | *p*-value | $\eta^2_p$ | $\beta$ (std) [95% CI] | *F*(df<sub>1</sub>, df<sub>2</sub>) | *p*-value | adj. *p*-value<sup>1</sup> | $\eta^2$ | $\beta$ (std) [95% CI] |
| **Age (years)** | *F*(1, 217) = 7.57 | .006 | 0.034 | 0.19 [+0.05, +0.32] | *F*(1, 247) = 7.72 | .006 | .038 | 0.030 | 0.17 [+0.05, +0.30] |
| Gender<sup>#</sup> | *F*(1, 216) = 0.01 | .930 | 0.000 |  | *F*(1, 245) = 0.23 | .634 | .836 | 0.001 |  |
| Occupation | *F*(4, 217) = 2.16 | .075 | 0.038 |  | *F*(4, 238) = 1.88 | .115 | .298 | 0.031 |  |
| Pre-existing health awareness | *F*(1, 217) = 1.48 | .225 | 0.007 | 0.08 [-0.05, +0.21] | *F*(1, 247) = 2.63 | .106 | .298 | 0.011 | 0.10 [-0.02, +0.23] |
| **Chronotype (MSF<sub>sc</sub>, h)** | *F*(1, 172) = 4.99 | .027 | 0.028 | -0.20 [-0.38, -0.02] | *F*(1, 202) = 9.41 | .002 | .032 | 0.044 | -0.21 [-0.35, -0.08] |
| Social jetlag (h) | *F*(1, 172) = 0.85 | .358 | 0.005 | 0.08 [-0.09, +0.26] | *F*(1, 212) = 1.48 | .226 | .489 | 0.007 | -0.08 [-0.22, +0.05] |
| Sleep duration (workdays, h) | *F*(1, 217) = 0.25 | .621 | 0.001 | -0.04 [-0.21, +0.12] | *F*(1, 246) = 0.08 | .772 | .836 | 0.000 | 0.02 [-0.11, +0.14] |
| Sleep duration (free days, h) | *F*(1, 217) = 1.22 | .271 | 0.006 | 0.09 [-0.07, +0.26] | *F*(1, 246) = 0.09 | .762 | .836 | 0.000 | 0.02 [-0.11, +0.14] |
| Shift work | *F*(1, 217) = 0.01 | .922 | 0.000 |  | *F*(1, 247) = 0.04 | .849 | .849 | 0.000 |  |
| State | *F*(5, 217) = 1.00 | .420 | 0.022 |  | *F*(6, 240) = 0.66 | .679 | .836 | 0.016 |  |
| Residence | *F*(2, 217) = 1.42 | .245 | 0.013 |  | *F*(2, 243) = 0.34 | .711 | .836 | 0.003 |  |
| Latitude (°) | *F*(1, 217) = 1.30 | .256 | 0.006 | -0.29 [-0.78, +0.21] | *F*(1, 245) = 2.66 | .104 | .298 | 0.011 | -0.10 [-0.23, +0.02] |
| Longitude (°) | *F*(1, 217) = 0.84 | .362 | 0.004 | 0.62 [-0.72, +1.96] | *F*(1, 245) = 0.57 | .452 | .836 | 0.002 | 0.05 [-0.08, +0.17] |

Effect sizes: partial $\eta^2$ (full model) and $\eta^2$ (= $R^2$ , univariate). Standardized coefficients ( $\beta$ ) are reported for continuous predictors only. <sup>#</sup>Estimated using the Male/Female subset only due to the small sample size of “other”. <sup>1</sup>adjusted for multiple comparisons using the Benjamini–Hochberg procedure. Chronotype and social jetlag were set to missing for shift/night workers, who lack a valid MSF<sub>sc</sub>, so their full-model estimates are based on non-shift respondents (n = 193), whereas all other predictors were estimated on the full control group.

**Table 3.** Moderation analysis of the health-risk messaging effect on attitudes toward DST.

| Moderator | *F*(df1, df2) | *p*-value | Conditional effect of condition on attitudes (simple slopes) |
| --- | --- | --- | --- |
| **Pre-existing health awareness** | *F*(1, 495) = 10.82 | .001 | Low (−1 SD): *B* = -0.32, *SE* = 0.16, *p* = .046; Mean: *B* = -0.70, *SE* = 0.11, *p* < .001; High (+1 SD): *B* = -1.08, *SE* = 0.16, *p* < .001 |
| Social jetlag (h) | *F*(1, 429) = 0.01 | .909 | Low (−1 SD): *B* = -0.70, *SE* = 0.18, *p* < .001; Mean: *B* = -0.71, *SE* = 0.12, *p* < .001; High (+1 SD): *B* = -0.72, *SE* = 0.18, *p* < .001 |
| State | *F*(5, 483) = 0.84 | .522 | NSW/ACT: *B* = -0.52, *SE* = 0.20, *p* = .009; VIC: *B* = -0.79, *SE* = 0.22, *p* < .001; QLD: *B* = -1.07, *SE* = 0.27, *p* < .001; other states ns |
| DST status | *F*(1, 492) = 1.37 | .242 | DST: *B* = -0.59, *SE* = 0.13, *p* < .001; No DST: *B* = -0.89, *SE* = 0.22, *p* < .001 |
| Residence | *F*(2, 489) = 0.32 | .727 | Urban: *B* = -0.77, *SE* = 0.14, *p* < .001; Rural: *B* = -0.68, *SE* = 0.63, *p* = .281; Suburban: *B* = -0.54, *SE* = 0.24, *p* = .026 |
| **Perceived credibility of information** | *F*(1, 495) = 54.96 | < .001 | Low (−1 SD): *B* = 0.08, *SE* = 0.16, *p* = .610; Mean: *B* = -0.74, *SE* = 0.11, *p* < .001; High (+1 SD): *B* = -1.55, *SE* = 0.15, *p* < .001 |
| ns indicates not significant. #Estimated using the Male/Female subset only due to the small sample size of “other”. |  |  |  |

### Data availability statement

The data and R code underlying this article have been deposited in the Open Science Framework (OSF) repository and are available at https://doi.org/10.17605/OSF.IO/KGX98.

## Results

### Participant characteristics of the study groups from the pre-registered survey on DST and health-risk messaging

To assess how and to what extent public attitudes toward DST, policy preferences, and its perceived health consequences (*e.g.,* beliefs about negative DST-related health effects and costs outweighing benefits) are influenced by exposure to DST-related health-risk information, we conducted a preregistered, hypothesis-driven online survey with 499 Australian adults, who were randomly assigned to receive either neutral information about DST (control condition; n = 249) or DST-related health-risk messaging (health-risk condition; n = 250). The preregistered design also tested whether individual differences, including demographics, pre-existing health awareness (*i.e.,* an individual’s general attentiveness to health-related topics), and perceived message credibility, moderate this effect. Because chronotype and social jetlag represent individual-level factors that may shape attitudes toward DST, we also assessed respondents’ chronobiological parameters using the µMCTQ, a validated questionnaire that provides estimates of chronotype, social jetlag, and sleep duration [38].

Of note, both groups were comparable in demographic, socioeconomic and geographic characteristics (see Table 1). More specifically, both groups had a balanced gender distribution, with slightly more women than men (control: 48% male, 51% female; health-risk: 43% male, 56% female), and a mean age of 38-39 years. Most respondents in both groups were employed in managerial, professional, technical, or scientific roles (control: 56%; health-risk: 57%), with others in clerical/secretarial (control: 16%; health-risk: 14%), sales/personal service (control: 12%; health-risk: 15%), or manual occupations (control: 15.6%; health-risk: 14.3%). Geographically, respondents from both groups were predominantly from DST-observing jurisdictions (control: 75%; health-risk: 69%) and mainly resided in urban areas (control: 74%; health-risk: 73%). Both groups also reported a moderately high level of pre-existing health awareness (control: M = 5.20, SD = 0.96; health-risk: M = 5.19, SD = 0.91) and thus general interest in health-related topics. Chronotype was ∼17 min later in the control group, a small baseline difference (SMD = 0.21).

### Baseline DST attitudes, health perceptions, and how they are moderated by individual differences without health-risk messaging

As a first step of our analysis, we examined responses from the control group only, representing unmanipulated public opinion regarding DST attitudes and health perceptions, to establish a baseline that is critical for contextualizing and interpreting subsequent analyses of the health-risk condition within a broader context. In the control condition, and thus without prior DST-related health-risk messaging, 59.8% of respondents expressed a generally positive attitude toward DST, with most falling in the moderate range (somewhat agree, 29.7%; agree, 25.3%; strongly agree, 4.8%). Opposition to DST was less common, reported by 20.1% of respondents overall, and tended to be moderate in strength (somewhat disagree, 16.9%; disagree, 2.0%; strongly disagree, 1.2%), and 20.1% were neutral (Fig. 1A, B). These findings suggest that, in the absence of DST-related health-risk messaging, public support for DST is moderately positive rather than strongly polarised. Preferences for adopting or maintaining DST policy at the state level followed a comparable pattern, with 57.3% indicating support and 31.3% opposition (Fig. 1C, D). In this case, however, responses were more polarised, with a greater proportion of respondents selecting the extreme categories of agreement and disagreement (Fig. 1B). This stronger polarisation may reflect the more concrete and policy-relevant framing of the question (*e.g.,* “I support eliminating/introducing daylight saving time in my state”), which could encourage firmer positions compared with the more general attitude questions.

By contrast, perceptions of DST’s health consequences indicated limited concern: only 20.5% endorsed the statements about adverse health effects (Agree [5–7]), whereas a majority rejected them (51.8%; Disagree [1–3]) and 27.7% were neutral (Fig. 1C). This result prompted us to examine how DST attitudes, policy support, and perceived health consequences were interrelated. We observed that a positive attitude toward DST was strongly correlated with support for DST policy (Fig. 1D; Spearman’s ρ = 0.88, n = 246, *p* < .001). In contrast, perceiving DST as harmful to health was associated with less favourable attitudes toward DST (Fig. 1E; Spearman’s ρ = -0.74, n = 249, *p* < .001) and with lower support for keeping or introducing DST policy (Fig. 1F; Spearman’s ρ = -0.58, n = 246, *p* < .001).

We then assessed which individual differences, including geographical location and sociodemographic factors, were associated with attitudes toward DST. Respondents had an average mid-sleep on free days corrected for sleep debt (MSF_sc_), a measure of chronotype, of 3.53 h (SD = 1.32, n = 204; Table 1), representing an intermediate chronotype. The average relative social jetlag was 0.51 h (SD = 0.68, n = 214), suggesting a relatively low circadian misalignment. We found that respondents with an older age (Fig. 2A; Table 2) and earlier chronotypes (*i.e.,* individuals with a low MSF_sc_; Fig. 2B; Table 2) expressed more favourable attitudes toward DST; both associations remained statistically significant after correction for multiple testing. None of the other tested factors significantly influenced attitudes toward DST (Table 2).

**Fig. 2.**
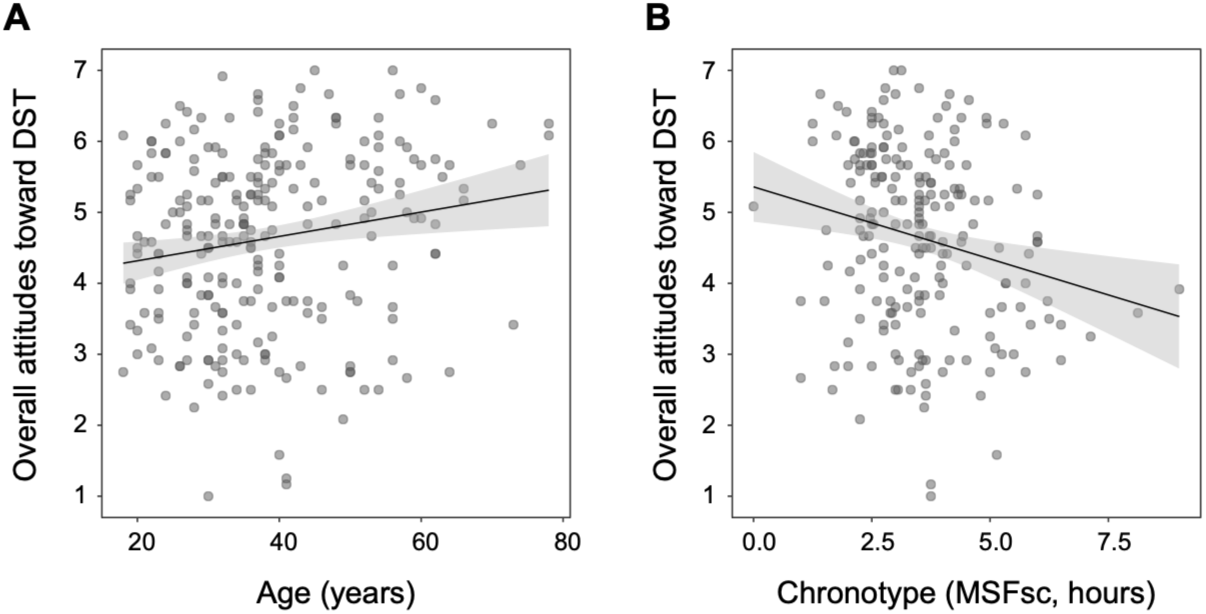
Influence of age and chronotype on overall attitudes toward DST in the absence of the health-risk message. **(A, B)** Univariate relationships (Table 2) between overall attitude toward DST and age (A) and chronotype (B) in the control group.

Together, these findings show that a modest majority of respondents in the control group support DST policy in Australia, with support closely linked to positive attitudes toward DST, lower perceptions of DST-related health consequences, and especially favourable views among older individuals and those with earlier chronotypes.

### How DST-related health-risk messaging shapes DST attitudes and perceived health consequences: moderating roles of pre-existing health awareness and message credibility

In the control condition, support for DST tended to align with lower concerns about its health consequences (Fig. 1E, F), consistent with our initial hypothesis that DST-related health-risk information has the potential to reduce support for DST. Next, we proceeded with the preregistered analysis examining how, and to what extent, exposure to health-risk information would modify attitudes and support for DST, and whether these effects varied with individual differences. Accordingly, we compared responses from the control and health-risk message conditions. Compared with the control condition, respondents in the health-risk condition who were exposed to DST-related health-risk information expressed less favourable overall attitudes toward DST (Fig. 1B; M = 3.93, SD = 1.31, n = 250 *vs*. control M = 4.63, SD = 1.26, n = 249, F(1,497) = 36.79, *p* < .001). They also reported a lower preference for adopting or maintaining DST policy (Fig. 1D; M = 3.76, SD = 2.04, n = 250 *vs.* control M = 4.45, SD = 2.09, n = 246, F(1,494) = 13.52, p < .001). Conversely, respondents in the health-risk condition perceived greater health consequences associated with DST (Fig. 1F; M = 4.70, SD = 1.35, n = 250 *vs.* control M = 3.52, SD = 1.31, n = 249, F(1,497) = 97.34, *p* < .001). When support was broken down by response category, exposure to the health-risk messaging reduced the proportion of respondents supporting DST (Fig. 1A; Agree [5–7]: 57.3% to 43.6%) and increased both opposition (Disagree [1–3]: 31.3% to 42.0%) and neutrality (Neutral [4]: 11.4% to 14.4%). Thus, health-risk messaging consistently reduced modestly favourable attitudes.

We next examined potential, preregistered moderators of this health-risk message effect on attitudes toward DST policy. We found that pre-existing health awareness moderated the impact of the health-risk message (Table 3). Individuals with higher general health awareness showed a stronger shift toward negative attitudes after receiving the health-risk information (Fig. 3A, B). Importantly, pre-existing health awareness was not associated with DST attitudes in the control group (Table 2), suggesting limited public knowledge of DST-related health risks and indicating that the observed effect reflected genuine message-driven change rather than baseline differences. In addition, perceived message credibility moderated the effect of the health-risk message on attitudes towards DST. The difference in attitudes toward DST between the health-risk and control conditions increased with perceived credibility: at low credibility (−1 SD), there was no detectable difference; at mean credibility, attitudes were lower in the health-risk group; at high credibility (+1 SD), the difference was largest (Fig. 3C, D; Table 3). Thus, message credibility plays a critical role in shaping how health-risk messaging influences attitudes toward DST.

**Fig. 3.**
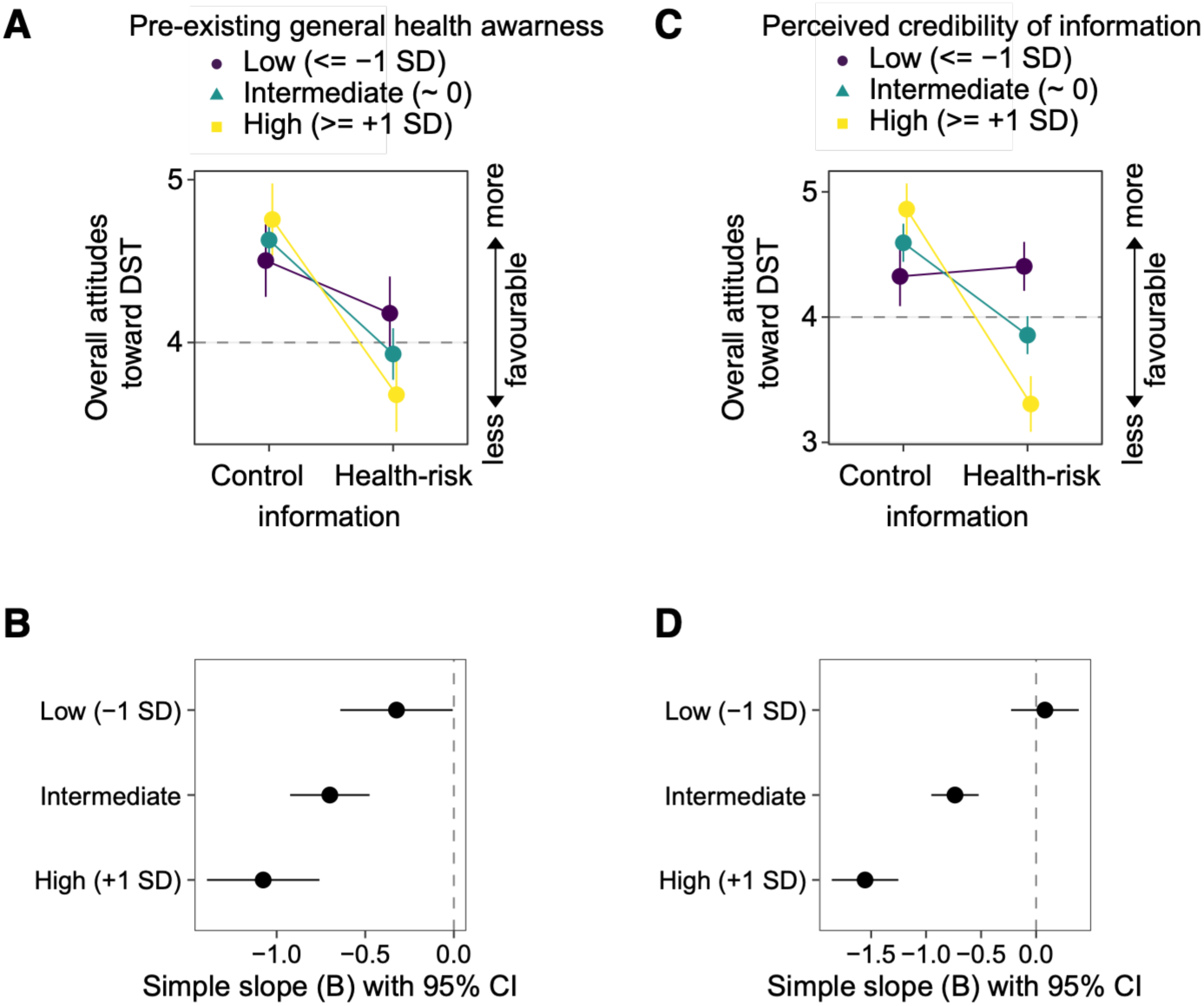
Moderation of health-risk messaging effects on DST attitudes by pre-existing health awareness and credibility of the information. **(A, C)** Interaction plots of estimated marginal means (EMMs) from a linear model with condition × moderator. Lines show predicted attitudes for control *vs.* health-risk information at low (−1 SD), intermediate, and high (+1 SD) levels of the moderator: pre-existing health awareness (A) and perceived message credibility (C). Error bars are 95% CIs; the dashed horizontal line marks the scale midpoint (y = 4). **(B, D)** Simple-slope forest plots. Points are unstandardized simple slopes (B) for the effect of condition (health-risk *vs*. control information) at low, intermediate, and high levels of the same moderators; horizontal bars are 95% CIs; the dashed vertical line marks B = 0.

Given the strong effects of DST-related health-risk messaging on attitudes toward DST and perceptions of its health consequences, we next tested whether the latter mediated the effect of messaging, and whether this pathway depended on the perceived credibility of the information provided (*i.e.*, a first-stage moderated mediation). As assumed, the health-risk message increased perceived negative health consequences (a = 1.25, SE = 0.11, *p* < .001; where higher scores indicate greater perceived consequences), and stronger perceived consequences in turn predicted lower support for DST (b = −0.79, SE = 0.03, *p* < .001). The indirect effect of condition on DST attitudes via perceived health consequences was significant (ab = −0.98, 95% CI [−1.17, −0.81]). A smaller but still significant direct effect of the health-risk message on attitudes also remained after accounting for perceived health consequences (c′ = +0.24, SE = 0.08, p = .001), indicating partial mediation. Importantly, the indirect pathway was moderated by perceived credibility: the effect of the health-risk message on perceived health consequences was small at low credibility (−1 SD: a = 0.32, SE = 0.16, *p* = .045), but became substantially stronger at high credibility (a = 2.18, SE = 0.16, *p* < .001 at +1 SD). As a result, the size of the indirect effect increased in magnitude with credibility, increasing from ab = −0.25 (95% CI [−0.50, −0.01]) at low credibility to ab = −1.72 (95% CI [−2.00, −1.44]) at high credibility. Together, these results show that increased perception of health risks was the primary mechanism through which health-framed messaging reduced DST support, and that this mechanism was stronger when the message was perceived as more credible.

## Discussion

Despite its societal relevance, public attitudes toward DST in Australia, and the individual differences in shaping them, remain insufficiently understood. More broadly, how awareness of DST-related health impacts influences these attitudes remains unclear, with important implications for evidence-based public health communication and policymaking within and beyond Australia. In this study, we observed that overall attitudes toward DST in Australia were closely associated with participants’ perceptions of its health consequences. We further demonstrated that DST-related health-risk information, particularly when regarded as credible, amplified concerns and shifted favourable views toward reduced support for DST policy.

In the control condition, slightly more than half of respondents expressed a positive attitude toward DST, while the remainder were neutral or opposed. When evaluating our results in direct comparison with those reported by a previous study [42], which found 73.3% (377/514) preference for DST in Eastern Australia (*i.e.,* Queensland, New South Wales, the Australian Capital Territory, Victoria, and Tasmania), we observed lower levels, with 52.0% (103/198) in our sample favouring keeping or introducing DST. This discrepancy may partly reflect differences in study design such as sampling strategies, question framing, and whether neutral opinions were permitted. It may also indicate that, in some states, support for DST may not have shifted substantially since the last state referenda (about 45.5% and 46% voted in favour in Queensland in 1992 and Western Australia in 2009, respectively [43]), and that a clear national consensus on DST policy has not yet emerged, a topic expected to be further addressed by upcoming investigations, such as the ongoing survey being conducted by the Sleep Health Foundation [44].

Regarding participant individual differences, we found that older participants and those with an earlier chronotype, and thus preference for earlier sleep-wake timing, expressed more favourable attitudes toward DST. By contrast, younger participants and those with later chronotypes, who have a later sleep-wake timing preference, were less favourable. This pattern is consistent with the well-documented prevalence of late chronotypes among adolescents [26,45,46] and with evidence that individuals with later chronotypes are more negatively affected by DST [19,30,47–50] and more likely to favour abolishing seasonal clock changes [51,52]. In addition, older participants’ more favourable views may also stem from greater temporal flexibility, for example due to retirement or reduced caregiving responsibilities [53,54], which lessens the practical disruption of DST. Consistent with this assumption, retirement has been linked to longer sleep, improved sleep quality, and reduced social-biological misalignment [55–58]. Since sleep health disparities disproportionately affect adolescents, later chronotypes, and individuals with early schedules [59,60], our observation that these groups express lower support for DST aligns with the view that DST policy has implications for social equality.

Moreover, our findings about how DST-related health-risk messaging shapes attitudes contribute to the broader debate on health communication and underscore the need for DST policy decisions to be guided by empirical evidence rather than individual beliefs or anecdotal accounts [1,17,36,37]. Yet, while the public views health as central to DST policymaking [52], health-related evidence remains largely absent from the policy debate [17,37]. For example, a recent analysis of US Senate debates about DST policy reported that only 15% of arguments were research-based, while 85% relied on non-research related justifications [37]. Here, we showed that positive attitudes toward DST, along with support for its adoption or continuation, are accompanied by a low perception of DST-related health risks. Importantly, however, exposure to health-risk messaging was a strong factor in shifting public opinion about DST. This shift was primarily driven by changes in how individuals perceive its health consequences, indicating a genuine interest in DST-related health-risk information. Of note, while DST-related health-risk messaging had the strongest effects on individuals with high health awareness, it consistently reduced support for DST across demographic and geographic subgroups, underscoring the central role of health-risk evaluations in shaping public support.

However, the shift in attitudes was strongly contingent on how credible the information was perceived to be, underscoring the role of message credibility in shaping public opinion. Although the health-risk information about DST was informed by statements issued by professional circadian and sleep societies and was generally viewed as credible by the participants, our findings showed a clear divide: participants who regarded the message as credible reduced their support for DST, whereas those who considered it less credible showed no such shift. More broadly, and in this context, our observations align with research in psychology, particularly on attitudes, persuasion, and health psychology, which emphasises the importance of message framing and survey design in shaping attitudes and behavioural intentions (*e.g.*, [61,62]). In addition to finding that perceptions of credibility shaped responses to DST-related health-risk information, we also observed that question framing influenced the extent to which participants expressed support for DST, with relatively moderate attitudes toward DST becoming more polarised when framed in terms of concrete policy actions such as abolishing or introducing DST. This divergence underscores how the framing of survey items, from general opinions to specific policy choices, can amplify attitude extremity. Therefore, public messaging about DST should consider not only the potential health consequences it highlights but also how the information is framed, as framing can shape support for policy change, as exemplified by research on climate change communication [63]. Taken together, even as the health effects of DST remain an active area of research and future studies may refine our understanding of the associated risks, our findings provide clear evidence that DST-related health-risk messaging reduces support for DST policy by heightening perceptions of its associated health risks. However, while this highlights that the public considers health risks in their policy preferences, the cognitive route through which this change occurred remains a matter of future research. Specifically, it is unclear whether participants are engaged in central, reflective processing of the health-risk information presented, or whether the attitude shift was driven by more peripheral, affective responses such as fear or heightened concern. This aligns with prominent models of persuasion and health communication, such as the Elaboration Likelihood Model [61], which distinguishes between rational, effortful persuasion and heuristic or emotion-driven processing.

## Data Availability

The data and R code underlying this article have been deposited in the Open Science Framework (OSF) repository.

https://doi.org/10.17605/OSF.IO/KGX98

## Acknowledgements

This work was supported by the Australian National Health and Medical Research Council (Investigator grant #2016334) and the Alzheimer’s Association Research Fellowship (#AARF- 22-917584), which funded the survey and provided salary support for BDW, and by the Australian National Health and Medical Research Council (Synergy grant #2019260) and the National Institute of Health (#R01AG078241), which funded salary support for MW and FG.

## Author contributions

Conceptualisation: MW, BDW

Methodology: MW, CVH, BDW

Investigation: MW, CVH, BDW

Formal analysis: MW, CVH, BDW

Writing - original draft: MW, BDW

Writing - review & editing: MW, CVH, FG, BDW

Funding acquisition: FG, BDW

Project administration: BDW

## Competing interests

The authors declare no competing interests.

## Notes

### Competing Interest Statement

The authors have declared no competing interest.

### Author Declarations

This study was approved by the University of Queensland Ethics Committee under approval number 2025/HE000340.

### Summary of Updates

This version incorporates refinements to the data-processing and analysis pipeline, with corresponding updates to the descriptive statistics, tables, and figures. The main conclusions are substantively unchanged. In addition, the chronotype-by-message moderation, which was not part of the preregistered analysis plan, has been removed from Table 3 and the associated figure.

